# Survival analysis in breast cancer using proteomic data from four independent datasets

**DOI:** 10.1101/2020.12.03.20242065

**Authors:** Ágnes Ősz, András Lánczky, Balázs Győrffy

**Author notes:** **CORRESPONDENCE** Balázs Győrffy MD PhD, Semmelweis University Dept. of Bioinformatics, Tűzoltó u. 7-9, H-1094, Budapest, Hungary.

## Abstract

Breast cancer clinical treatment selection is based on the immunohistochemical determination of four proteins: ESR1, PGR, HER2, and MKI67. Our aim was to correlate immunohistochemical results to proteome-level technologies in measuring the expression of these markers. We also aimed to integrate available proteome-level breast cancer datasets to identify and validate new prognostic biomarker candidates.

We identified protein studies involving breast cancer patient cohorts with published survival and proteomic information. Immunohistochemistry and proteomic technologies were compared using the Mann-Whitney test. Receiver operating characteristics (ROC) curves were generated to validate discriminative power. Cox regression and Kaplan-Meier survival analysiss were calculated to assess prognostic power. The false discovery rate was computed to correct for multiple hypothesis testing.

The complete database contains protein expression data and survival information from four independent cohorts for 1,229 breast cancer patients. In all four studies combined, a total of 7,342 unique proteins were identified, and 1,417 of these were identified in at least three datasets. ESR1, PGR, HER2 protein expression levels determined by RPPA or LC-MS/MS methods showed a significant correlation with the levels determined by immunohistochemistry (p<0.0001). PGR and ESR1 levels showed a moderate correlation (correlation coefficient=0.17, p=0.0399). A panel of candidate proteins, including apoptosis-related proteins (BCL2,), adhesion markers (CDH1, CLDN3, CLDN7) and basal markers (cytokeratins), were validated as prognostic biomarkers. We expanded our established web tool to validate survival-associated biomarkers to include the proteomic datasets analyzed in this study (https://kmplot.com/analysis/).

Large proteomic studies now provide sufficient data enabling the validation and ranking of new protein biomarkers.

## INTRODUCTION

Breast cancer is one of the most frequently diagnosed cancers and the leading cause of cancer-related death in women (1). Routine utilization of histopathological markers has led to better survival outcomes in personalized therapy, while multigenic genomic and transcriptomic analyses have further defined clinically meaningful molecular subtypes (2). Genomics provides the ‘blueprint’ for cellular structure and functions, but due to its nature, it is always static, and the genome itself does not define the biological function. On the other hand, proteomics can show the physical structure of the cell, revealing a dynamic picture of active key functional elements. Proteomics can display the status of over 500,000 gene products defined by only approximately 30,000 genes. Overall, proteomics can provide a snapshot of the biological functions within a cancer cell. However, the availability of proteomic data derived from large patient cohorts is still limited.

Routine methods used for protein quantification include antibody-based techniques, such as immunohistochemistry (IHC) and reverse-phase protein array (RPPA), enzyme-linked immunosorbent assays (ELISA) and mass spectrometry (MS)-based technologies. ELISA invented in the 1970s is extensively used in laboratory practice for analyzing a small number of proteins, but its limitations in multiplexing requiring high developmental costs and well-characterized antibodies prevented its large-scale application (3). IHC is currently the gold standard method in routine pathological diagnosis, including the semiquantitative determination of ESR1, PGR and HER2 receptor status in breast tumors. Multiplexing of IHC is achieved in tissue microarrays, but even these achieve higher output by multiplexing the patient samples and not by multiplexing the proteins simultaneously evaluated. Nevertheless, tissue microarrays play a solid role in uncovering new biomarkers in cancer research (4). Although IHC is the most frequently used protein analysis method in oncology, it also has limits in the quantification and detection of activated proteins (5).

In contrast to antibody-based methods, the RPPA technique, introduced in 2001, immobilizes the whole protein lysate on a solid phase in multiple dots. A specific antibody solution is added to each array spot separately to achieve sensitive and simultaneous detection of proteins in small sample amounts (e.g., biopsy). RPPA requires well-specified antibodies, but it also makes it feasible to quantify the phosphorylation status of proteins and thus enables the characterization of entire pathways (6).

Mass spectrometry (MS)-based technologies have rapidly advanced in recent years. In addition to speed, the second most prominent advantage of these methods is their ability to facilitate de novo identification and quantification of multiple proteins simultaneously. However, MS requires high initial cost, manual and time-consuming sample preparation, and an experienced technician to run the samples and interpret the data (7). Three major quantitative MS-based techniques have been developed: shotgun (or discovery), directed, and targeted proteomics. The shotgun method is based on the sequencing of peptides digested from the whole proteome and analyzing them via liquid chromatography and tandem mass spectrometry (LC-MS/MS) and automated database searching (8). Then, the protein quantity is calculated from the signal of detected peptides (ion intensity) or recorded number of MS/MS spectra (spectral counting). Protein abundance is normalized to the background proteome signal of measured samples (LFQ) or to an internal standard added to a labeled experiment (9, 10).

These methods enable comprehensive large-scale analysis of the human proteome. International initiatives have emerged to facilitate collaboration and data sharing. The Human Proteome Organization (HUPO, www.hupo.org) initiated in 2010 the Human Proteome Project (HPP) aiming for the determination of the human proteome using a standardized analytical pipeline (11). Major data repositories for MS-based protein datasets include the ProteomeXchange Consortium (http://www.proteomexchange.org), PRIDE (http://www.ebi.ac.uk/pride), and PeptideAtlas (http://www.peptideatlas.org) (12). The Human Protein Atlas portal (www.proteinatlas.org) provides antibody-based data of normal and cancerous tissues (13). The Clinical Proteomic Tumor Analysis Consortium (CPTAC, https://cptac-data-portal.georgetown.edu/cptacPublic) of the National Cancer Institute curates combined genomic and proteomic data of multiple tumor types (14). Finally, a side project of The Cancer Genome Atlas (TCGA) Project, The Cancer Proteome Atlas (TCPA, https://tcpaportal.org/tcpa/index.html) contains a large RPPA-based protein expression cohort (9).

Breast cancer is classified into four molecular subtypes, each having different molecular and prognostic characteristics. In the clinical routine, immunohistochemistry is used to measure the presence of estrogen receptor (ESR1), progesterone receptor (PGR), human epidermal growth factor receptor 2 (HER2) and the proliferation marker MKI67. Evaluation of these biomarkers is mandatory to assign patients into clinically effective treatment subtypes termed basal (receptor negative), luminal A (ESR1 and PGR positive and low MKI67), luminal B (ESR1 and PGR positive and high MKI67), and HER2-enriched (HER2 positive ESR1 negative) (15). Of note, additional markers, including androgen receptor (AR), epidermal growth factor receptor (EGFR) and cytokeratins (CK), have also been proposed for biomarker-based subtyping (16, 17).

Proteomic datasets comprise a large amount of protein-level data for each included specimen, and therefore, these datasets can provide an opportunity to validate existing prognostic biomarkers. In addition, by simultaneously analyzing multiple proteins in the same sample cohort, one can compare and rank new biomarker candidates. However, utilization of these sample cohorts is difficult due to limited/unavailable clinical data, ambiguous analysis pipelines, and discrepant gene annotations. Here, our first goal was to establish a breast cancer proteomic resource database by combining samples from multiple large independent studies. Then, we aimed to utilize this resource to validate and rank prognostic protein biomarkers in breast cancer.

## MATERIAL AND METHODS

### Construction of the integrated protein database

We searched for publications and datasets containing proteome and survival data for breast cancer patients in PubMed, The Cancer Proteome Atlas (TCPA) (9) and the ProteomeXchange Consortium (18) portals. The search terms ‘human’, ‘breast’, and ‘cancer’ were used to identify eligible datasets. Only studies with available protein expression data generated by either mass spectrometry or RPPA, clinical survival information, and at least 50 cancer patients met our inclusion criteria. Four protein datasets met these conditions (9, 19-21). Due to the use of different platforms and analysis methods, it was not possible to merge the datasets into a single unified dataset. Therefore, each dataset was processed separately. In the analyses, the author-reported normalized expression data were used. **Figure 1** summarizes the pipeline of data filtering.

**Figure 1.**
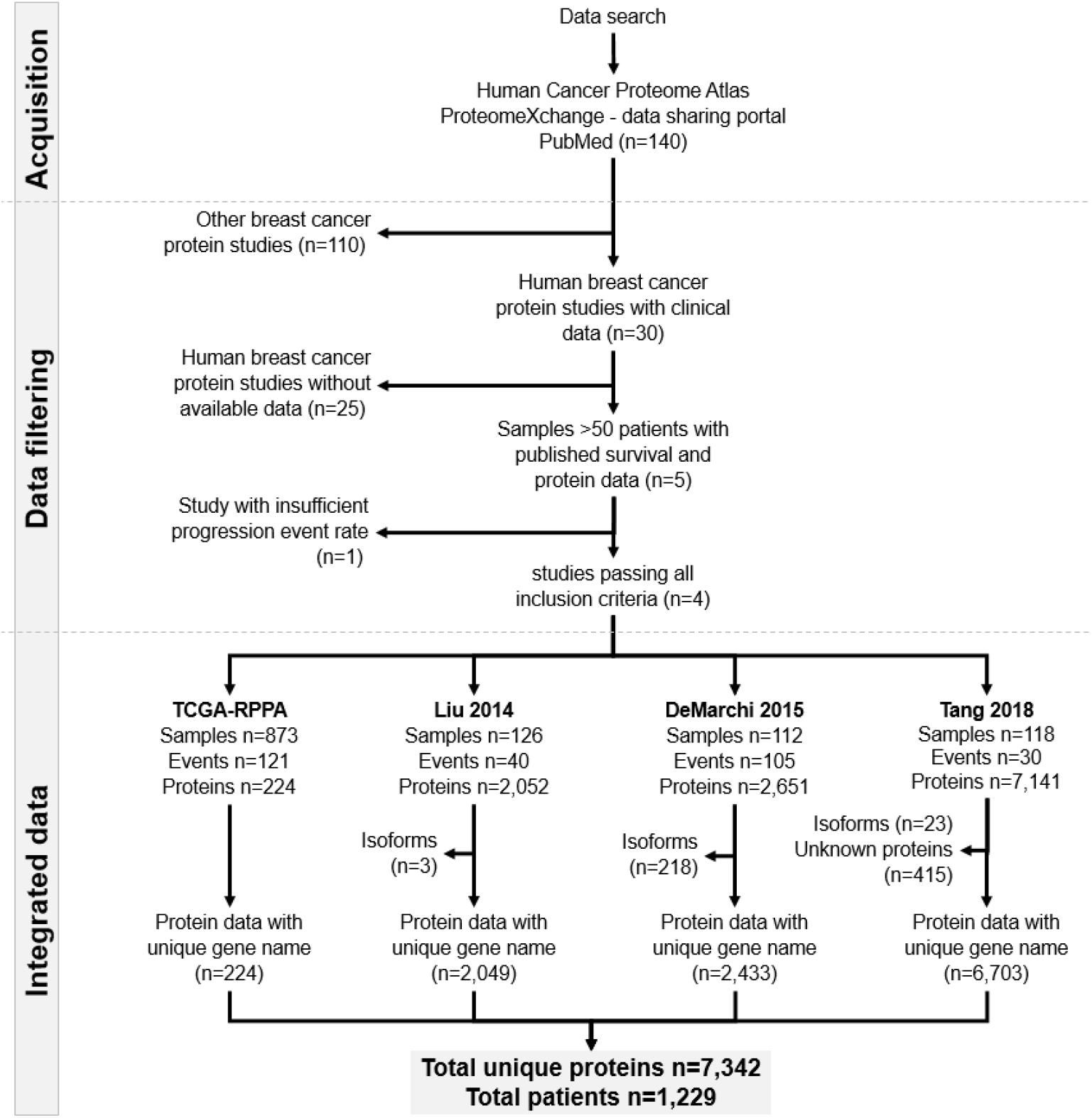
Data acquisition workflow, the number of samples and unique proteins in each included dataset.

### Protein annotation

In each dataset, the protein annotation generated by the authors was the starting point and duplicated and non-annotated proteins were removed. In addition, UniProt IDs were used to identify gene symbols corresponding to the same genes. The final integrated table of all annotated proteins in the database, including the gene symbol, UniProt ID and TCPA antibody list, is provided as **Supplemental Table 1**.

### Validation of proteome-based protein level determination

To determine how effective recent proteomic technologies are in assessing the actual protein levels, we compared proteome-based results to conventional immunohistochemistry results. Such data were available in multiple data sets for genes with therapeutic importance, including ESR1, PGR, HER2 and MKI67. All validation analyses were performed in each of the four cohorts separately. In the case of MKI67, we also compared the expression between normal and tumor tissue, as this was available in one dataset.

### Correlation between protein biomarker candidates and survival

We performed a PubMed search to identify previously published biomarker candidates related to survival using the search terms ‘breast cancer’, ‘protein’, ‘cohort’, ‘marker’, and ‘survival’. Publications describing cell lines, other tumor types, those not investigating a tumor tissue, and studies with fewer than 100 patients were excluded. After these restrictions, 53 publications remained. In addition, we examined ten additional publications describing breast cancer guidelines. In all 63 publications, a total of 91 proteins were described, 57 of which were present in our database. This list includes FDA-approved biomarkers, growth factor receptors, immune receptor ligands, basal and adhesion markers (cytokeratins, cadherins, and claudins), stem cell markers, and apoptotic markers **(Supplemental Table 2)**. We analyzed all 57 protein biomarkers used in breast cancer diagnostics for their prognostic power. The validation of the markers was performed separately in each dataset using overall survival and relapse-free survival time.

### Statistical analyses

Differential expression was evaluated using the Mann-Whitney test. Receiver operating characteristics (ROC) were computed to measure sensitivity and specificity and to validate discriminative power. ROC was also utilized to determine the optimal cutoff values to define cohorts based on the expression of the investigated proteins. Spearman rank correlation coefficients were calculated to assess the correlation of continuous variables. To measure the association between protein expression and survival length, the patients were grouped into high and low expression groups based on the expression of the selected protein. Then, the two groups were compared by Cox proportional hazards regression, and hazard ratios (HRs), 95% confidence intervals (CIs) and log-rank p values were calculated. Finally, for a selected set of markers, Kaplan-Meier plots were generated to display the different survival characteristics of the two cohorts. For cutoff values, each potential threshold was analyzed between the lower and upper quartiles, and the false discovery rate (FDR) was computed to correct for multiple hypothesis testing. The results were accepted as significant when p<0.05 and FDR<0.2.

### Survival analysis web tool

We previously created an online analysis platform utilizing transcriptome-level mRNA expression (22) and miRNA expression (23) data together with clinical, follow-up, and pathological data to assess the correlation between gene expression and survival in breast cancer. Here, we have established a new subsystem of this analysis platform. The complete proteomic database is now integrated into this system, and new biomarker candidates, as well as each biomarker assessed here, can be rapidly evaluated using the registration-free analysis site. In the tool, selection of the proteins can be performed using the gene symbol, the UniProt ID or the RPPA antibody name (https://kmplot.com/analysis/).

## RESULTS

### Integrated breast cancer protein database

Altogether, 140 datasets were identified, of which 30 studies had at least some clinical information for the included patients. We listed all these datasets in **Table 1**. After exclusion of those without survival data and other ineligible studies, four independent projects remained. These four datasets comprise 1,229 specimens and 7,342 unique proteins. The entire set of patients included 1,064 overall survival (OS) and 998 relapse-free survival (RFS) records. Two datasets had either only overall (Tang 2018) or relapse-free survival data (DeMarchi 2015). Median OS and RFS times varied between 27.6-96.5 months and 9.6-85.5 months, respectively. The mean age of the patients was 57.7±13.6 years. In line with our expectations, estrogen receptor-positive (ESR1+) patients represented approximately 67% of all samples, and almost half of the patients had nodal involvement (46%). Of note, the Liu 2014 dataset included triple negative breast cancer (TNBC), lymph node negative and treatment naive patients only. In the other studies, hormone therapy, primarily tamoxifen, was applied (59%). **Table 2** contains detailed clinical parameters for each included dataset used, and **Figure 2** shows selected clinical characteristics for these datasets.

**Table 1.** Overview of breast cancer proteomic studies.

| Reference | ProteomeXchange<br>/CPTAC ID | Method used | Survival | Sample<br>n | Protein n | Reason for<br>exclusion | Eligible |
| --- | --- | --- | --- | --- | --- | --- | --- |
| Tang et al. (2018) | PXD005692 | LC-MS/MS | available | 65 | 7141 | - | yes |
| Terunuma et al. (2014) | NA | GC-MS, LC-MS | available | 67 | NA | no protein data | no |
| Mertins et al. (2016) | S039 (CPTAC) | LC-MS/MS | available | 105 | 15369 | only 13 events | no |
| Huang et al. (2017) | S032 (CPTAC) | LC-MS/MS | not available | 24 | 12794 | no survival data | no |
| Waldemarson et al. (2016) | PXD000944 | 2D-DIGE, LC-MS/MS | available | 38 | 14000 | only 38 samples | no |
| Cifani et al. (2015) | PXD000691 | 2D-DIGE, LC-MS/MS | available | 38 | 3681 | only 38 samples | no |
| Liu et al. (2014)a | PXD000260 | nLC-MS/MS | available | 126 | 5000+ | - | yes |
| Liu et al. (2014)b | PXD000260 | nLC-MS/MS | available | 126 | 5000+ | - | yes |
| TCGA (2012) | NA | RPPA | available | 348 | 171 | - | yes |
| Bouchal et al. (2015) | PXD000029 | iTRAQ-2DLC-MS/MS | not available | 96 | 4405 | no survival data | no |
| Sjöström et al. (2015) | PXD001685 | LC-MS/MS; LC-SRM | not available | 80 | 778 | no survival data | no |
| De Marchi et al. (2015) | PXD000485 | LC-MS/MS | available | 112 | 3109 | - | yes |
| De Marchi et al. (2016) | PXD002381 | LC-MS/MS | not available | 38 | 3404 | no survival data | no |
| De Marchi et al. (2016) | PXD002381 | LC-MS/MS | not available | 38 | 4 | no survival data | no |
| Pozniak et al. (2016) | PXD000815 | LC-MS/MS | not available | 44 | 10124 | no survival data | no |
| Pedersen et al. (2017) | PXD005544 | TMT-HILIC; LC-MS/MS | not available | 34 | 4163 | no survival data | no |
| Zagorec et al. (2018) | PXD008012 | Ti(IV)-IMAC; LC-MS/MS | not available | 34 | 9000+ | no survival data | no |
| Tyanova et al. (2016) | PXD002619 | LC-MS/MS | not available | 40 | 10135 | no survival data | no |
| Jiang et al. (2015) | PXD002208 | LC-MS/MS | not available | 53 | 115 | no survival data | no |
| Haukaas et al. (2015) | NA | RPPA | not available | 191 | 150 | no survival data | no |
| Ternette et al. (2018) | PXD009738 | nUPLC-MS/MS | not available | 11 | 6275 | no survival data | no |
| Chen et al. (2018) | PXD007217 | LC-MS/MS | not available | 10 | 388 | no survival data | no |
| Naba et al. (2017) | PXD005554 | LC-MS/MS | not available | 4 | 1000 | no survival data | no |
| Gajbhiye et al. (2017) | PXD006441 | iTRAQ-SCX; LC-MS/MS | not available | 76 | 365 | no survival data | no |
| Chen et al. (2018) | PXD007572 | LC-MS/MS | not available | 56 | 556 | no survival data | no |
| Chen et al. (2017) | PXD005214 | LC-MS/MS | not available | 36 | 2413 | no survival data | no |
| Lobo et al. (2017) | PXD003106 | LC/MS-MS | not available | 40 | 4175 | no survival data | no |
| Braakman et al. (2017) | PXD003632 | nLC/MS-MS | not available | 38 | 2995 | no survival data | no |
| Muraoka et al. 2013 | PXD000066 | nLC-MS/MS | not available | 18 | 7092 | no survival data | no |
| Jordan et al. (2016) | PXD003322 | SPS-based MS3 | not available | 3 | 6300+ | no survival data | no |

**Table 2.** Detailed clinical features of the four protein datasets eligible for this analysis.

| Dataset<br>(Reference) | Platform<br>(Company) | Technology | Sample<br>size | Median<br>follow-up<br>(OS,<br>months) | Progression<br>events<br>(OS) | Median<br>follow-up<br>(RFS,<br>months) | Progression<br>events<br>(RFS) | ESR1+<br>(*) | PGR+<br>(*) | HER2+<br>(*) | Stage<br>(1/2/3/4) | Grade<br>(1/2/3) | Lymph-<br>node<br>positive | Age | Radiation<br>therapy | Hormone<br>therapy | Chemo-<br>therapy |
| --- | --- | --- | --- | --- | --- | --- | --- | --- | --- | --- | --- | --- | --- | --- | --- | --- | --- |
| TCGA-<br>RPPA<br>(9, 38) | 2470 Arrayer<br>(Quanterix) | RPPA | 873 | 27.6 | 121 | 25.3 | 64 | 627 | 532 | 133 | 128/505/<br>207/18 | - | 452 | 58.2±13.3 | 53 | 422 | 488 |
| Liu 2014<br>(19) | LTQ-Orbitrap-XL MS system<br>(ThermoElectron) | LC-MS/MS | 126 | 96.5 | 40 | 85.5 | 50 | 0 | 0 | 0 | - | 2/16/87 | 0 | 53.9±13.8 | - | 0 | 0 |
| DeMarchi<br>2015<br>(20) | LTQ-Orbitrap-XL MS system<br>(ThermoElectron) | LC-MS/MS | 112 | - | - | 9.6 | 105 | 112 | - | - | - | - | 104 | 61.1±11.2 | - | 112 | - |
| Tang 2018<br>(21) | LTQ MS system<br>(Thermo Fisher<br>Scientific) | LC-MS/MS | 118 | 50.0 | 30 | - | - | 32 | - | - | 6/46/13/0 | 8/19/28 | 27 | 54.5±15.7 | - | - | - |

OS: overall survival, RFS: relapse-free survival
\*ER, PGR, HER2 receptor status was identified using both gene expression and immunohistochemistry data in each cohort.

**Figure 2.**
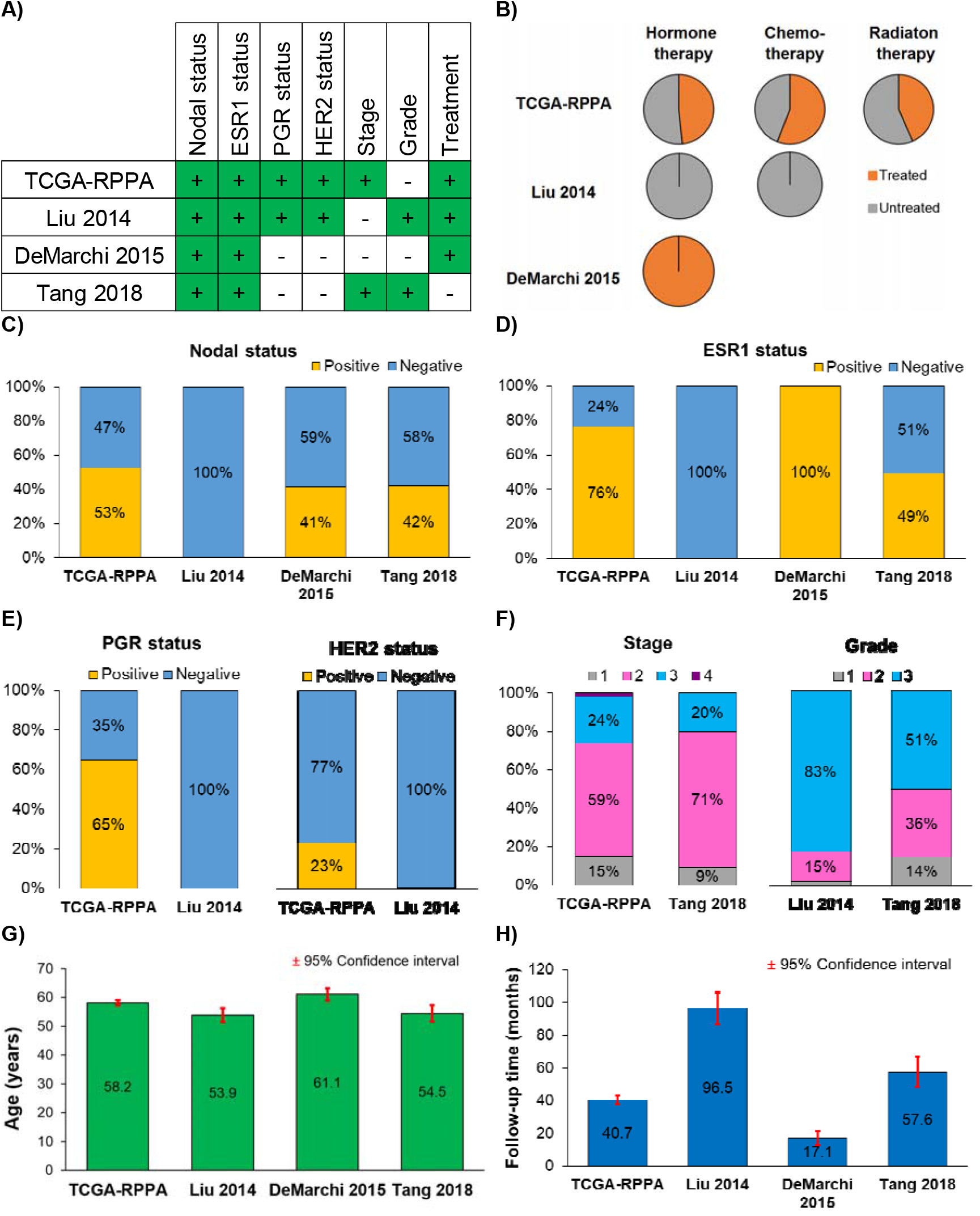
Clinical characteristics of the breast cancer patients used in this study. **A)** Availability of clinical data in the included cohorts; **B)** the proportion of patients treated with radiation, hormones or chemotherapy. **C)** Percentage of patients by nodal status in each dataset; **D)-E)** the proportion of patients by receptor status for ESR1, PGR and HER2 in each dataset; **F)** the distribution of stage and grade; **G)** the mean age of patients; and **H)** the mean follow-up time in each dataset.

The dataset generated using RPPA contains most of the patients (n=873) but least of the proteins (n=224). The other three datasets have combined >7000 protein records measured by LC-MS/MS technology. **Figure 3A** shows the proportions of detected proteins in each dataset combination. Only 39 proteins were measured in all datasets, while 1,356 overlapping proteins were evaluated in the three LC-MS/MS studies. A total of 4,731 proteins were detected in only one study, and most of them came from the Tang 2018 cohort (n=4,225). When mapping the measured proteins to cellular locations, the majority of proteins originated from the cytoplasm (36.3%), nucleus (32.2%) and cytosol (27.6%) (**Figure 3B** and **3C**). **Supplemental Table 1** includes all proteins.

**Figure 3.**
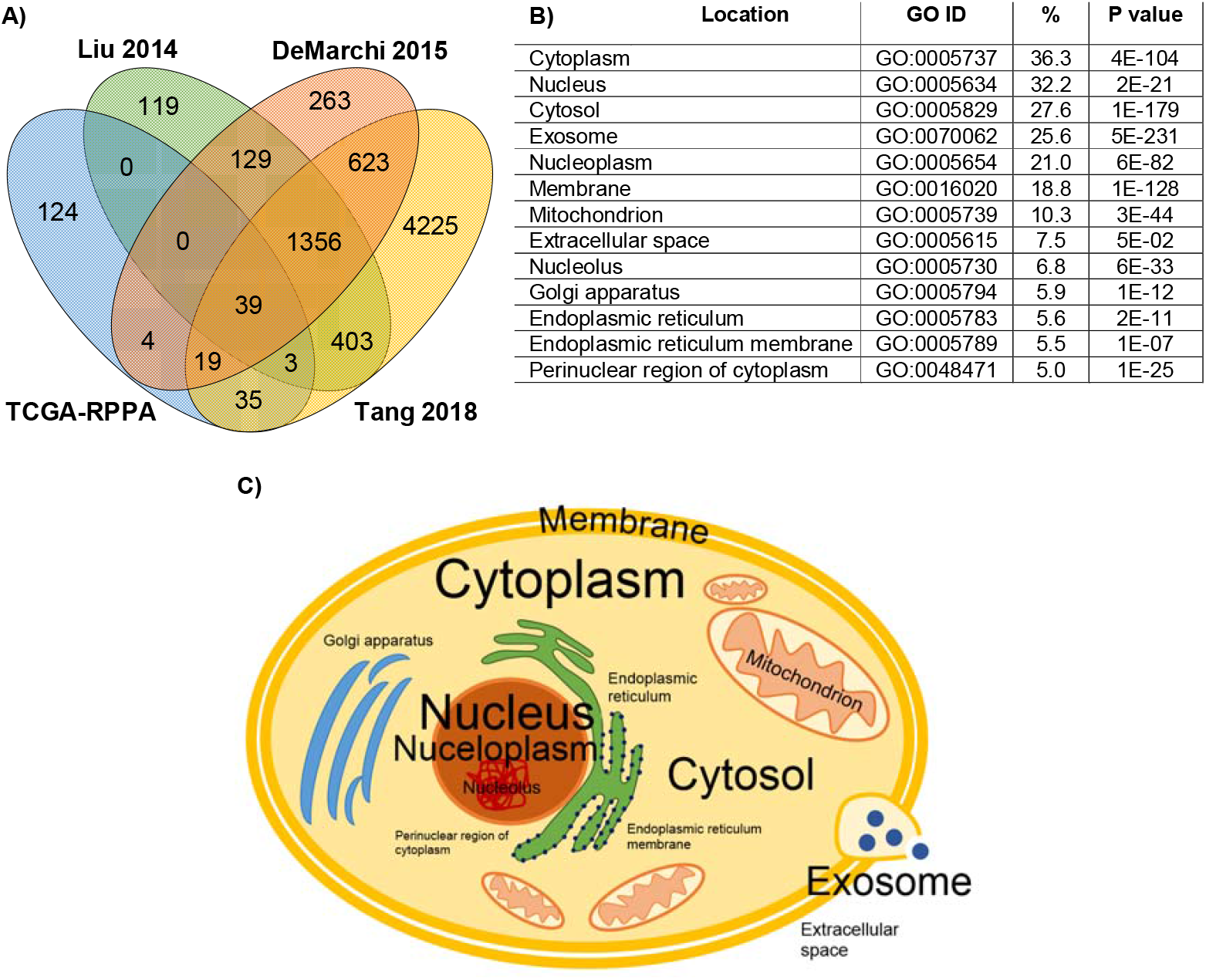
Proteins measured in multiple studies and their cellular localizations. **A)** Number of proteins represented in one, two, three, or four datasets, **B)** proportion of proteins present in various cellular components, and **C)** graphical representation of cellular origin of the analyzed proteins, where font size is relative to the proportion of proteins from that compartment.

### Evaluation of routine diagnostic biomarkers

ESR1, PGR and HER2 protein expression levels determined by RPPA were compared to IHC-based receptor status and the results revealed that protein expression and receptor status were highly significantly correlated with one another (p<0.0001) (see **Figure 4A-C)**. When running ROC analysis using RPPA-based continuous HER2 levels, the proteomic measurements delivered a substantial area under the ROC curve (AUC) of 0.74 (p=1.9e-20). ESR1 protein expression determined by LC-MS/MS also delivered a reliable correlation to IHC results (p=0.0423) **(Figure 4D)**. The AUC value for ESR1 levels determined by LC-MS/MS was 0.61 (p=0.03).

**Figure 4.**
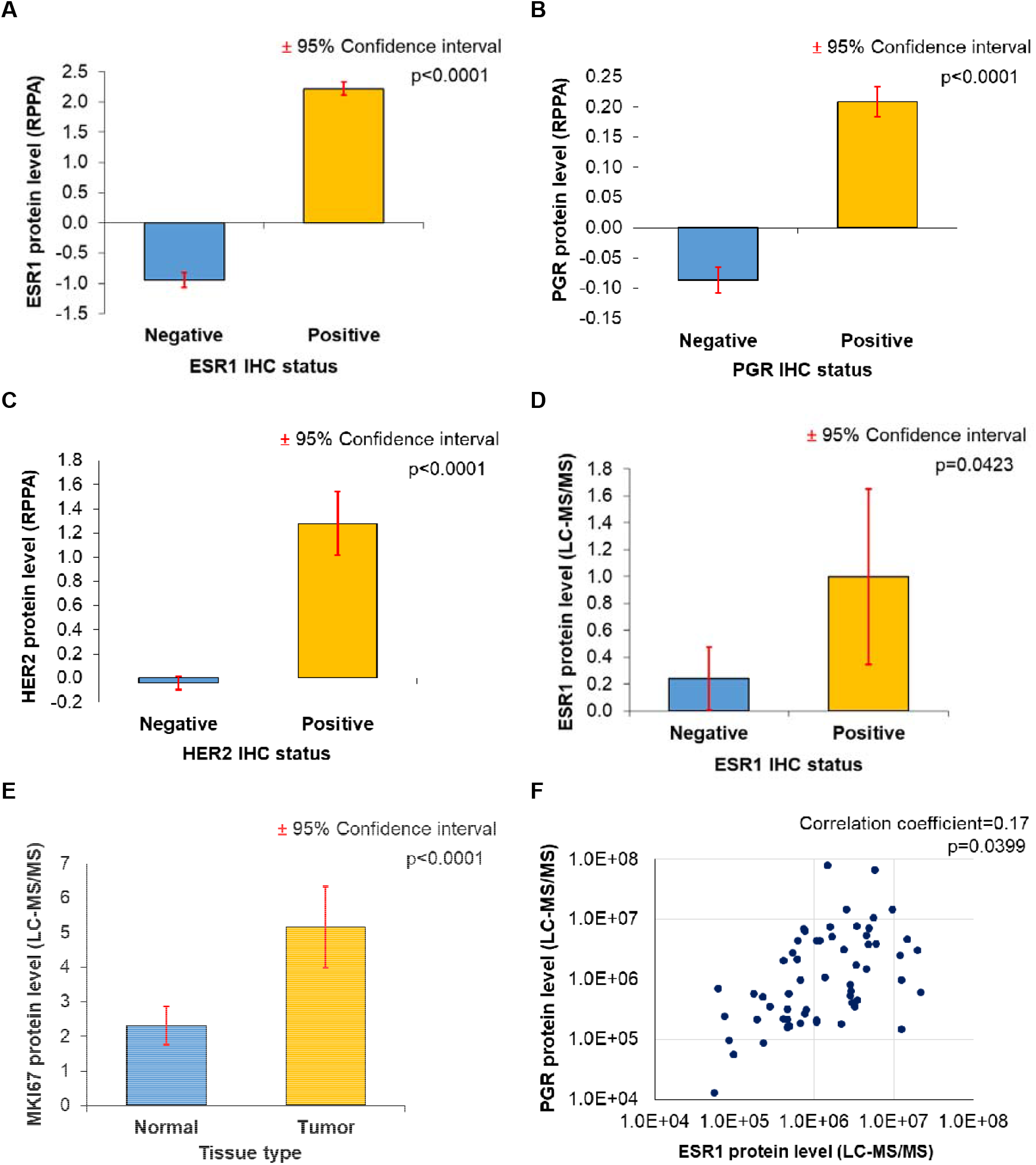
Validation of proteome-based molecular biomarker determination by comparing the results to those achieved by IHC-based receptor status determination. **A)-C)** ESR1, PGR, HER2 protein expression results determined by RPPA showed a significant correlation with IHC results. **D)** The correlation between ESR1 status by IHC and ESR1 protein expression levels measured by LC-MS/MS. **E)** MKI67 levels measured by LC-MS/MS showed higher expression in tumors than in normal samples in the Tang 2018 dataset (n=53). **F)** Correlation between ESR1 and PGR protein expression levels in LC-MS/MS data.

The Tang et al dataset included paired normal and tumor samples for 53 patients. When comparing the expression of the proliferation marker MKI67 between the normal and cancer samples, the tumor samples had significantly higher expression (fold change=2.22, p=0.0001) **(Figure 4E)**.

Finally, we also assessed the correlation between ESR1 and the ESR1-regulated gene PGR. In this analysis, we uncovered a moderate correlation between ESR1 and PGR protein expression levels, as determined by LC-MS/MS (correlation coefficient=0.17, p=0.0399, **Figure 4F**). Unfortunately, due to the limited availability of simultaneously collected data, it was not possible to analyze all possible clinical scenarios and to model molecular subtype determination based on proteomic datasets.

### Proteins with significant prognostic power

We assessed the link between survival and the expression of 63 proteins and their phosphorylated forms to validate their prognostic relevance in breast cancer **(Supplemental Table 2)**. The expression of 33 of 63 proteins had a significant correlation with patient outcome. Twelve proteins associated with OS only, nine proteins associated with RFS only, and twelve proteins (PGR, CDH1, BCL2, NDRG1, CTNNB1, APOD, PARP1, RBM3 and four cytokeratins: KRT18, KRT5, KRT6B, KRT17) were prognostic for both RFS and OS. Of these, three proteins (KRT18, APOD and CDH1) and four proteins (PGR, CDH1, CTNNB1, and BCL2) were confirmed to be related to OS and RFS, respectively, in at least two independent datasets. The results of the survival analysis for each of these proteins in terms of OS and RFS are displayed in **Table 3A** and **3B**, respectively.

**Table 3.**
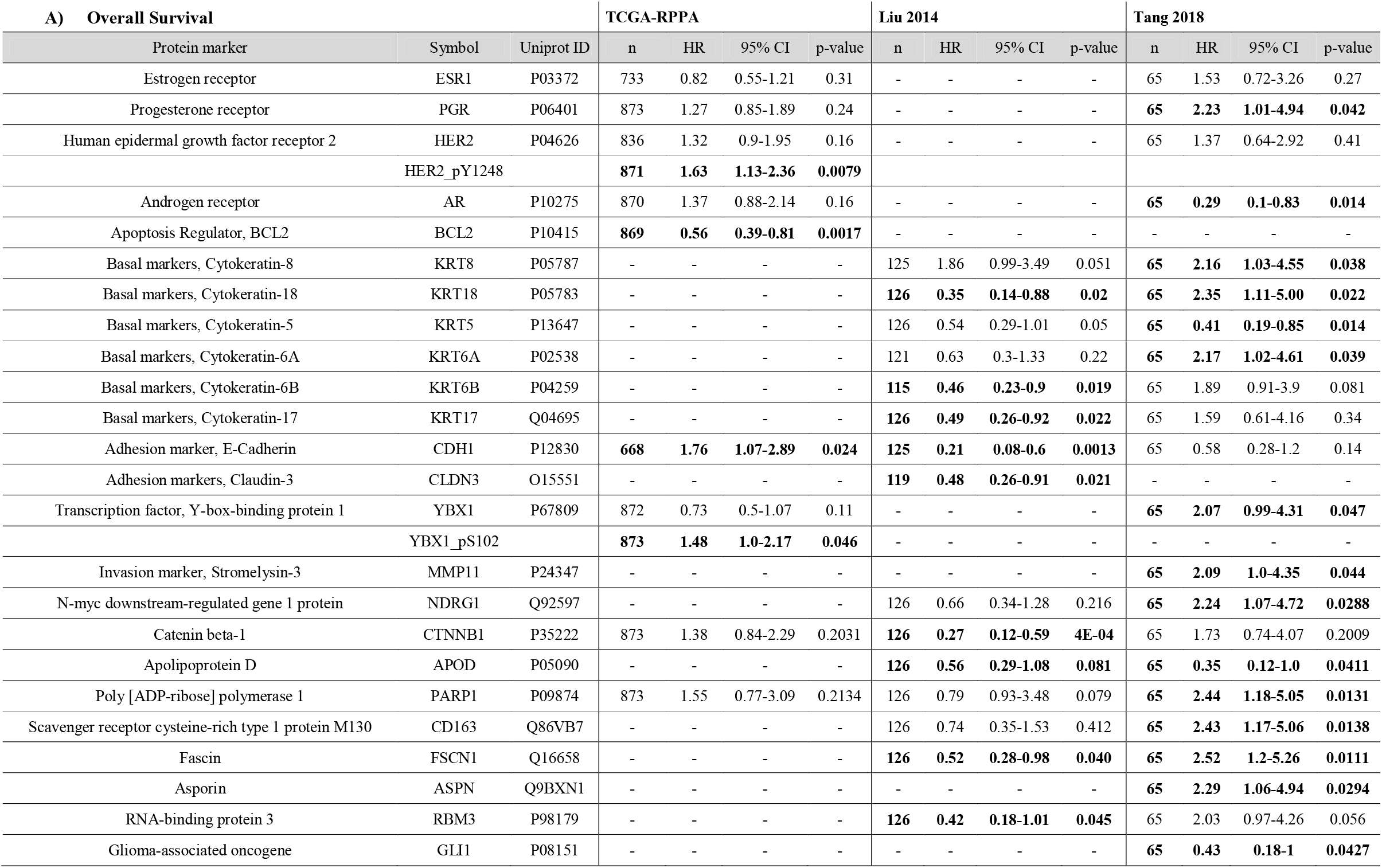
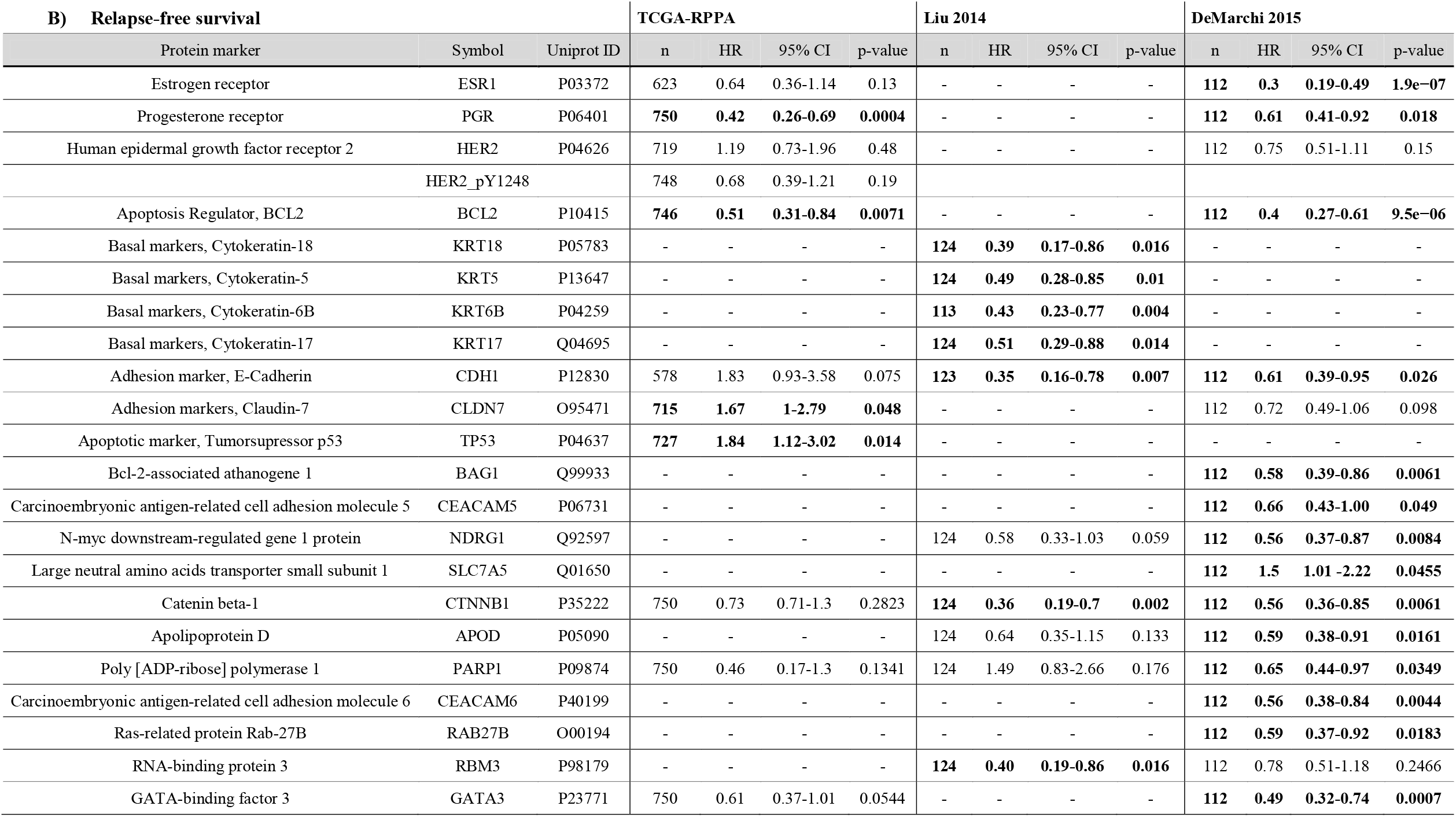
Protein markers with validated prognostic value in breast cancer when assessing the correlation between expression level and overall survival **(A)** and relapse-free survival **(B)**. *Bold: significant at p<0*.*05*.

A better overall survival outcome was associated with higher expression of E-cadherin (HR=0.21, 95%CI=0.08−0.6, p=0.0013) and the apoptosis regulator protein BCL2 (HR=0.6, 95%CI=0.39−0.81, p=0.0017). Higher BCL2 was also strongly related to longer relapse-free survival (HR=0.4, 95%CI=0.27−0.61, p=9.5e−06). While we also validated the prognostic value of the expression level of tyrosine 1248-phosphorylated HER-2 (HER2_pY1248) (HR=1.63, 95%CI=1.13−2.36, p=0.0079) using RPPA data, the expression level of nonphosphorylated HER-2 did not have a significant correlation with survival in any of the included datasets. Both estrogen receptor and progesterone receptor were linked to improved relapse-free survival (HR=0.3, 95%CI=0.19−0.49, p=1.9e−07 and HR=0.4, 95%CI=0.26−0.69, p=0.0004, respectively). Kaplan-Meier curves for these proteins are shown in **Figure 5A-F**.

**Figure 5.**
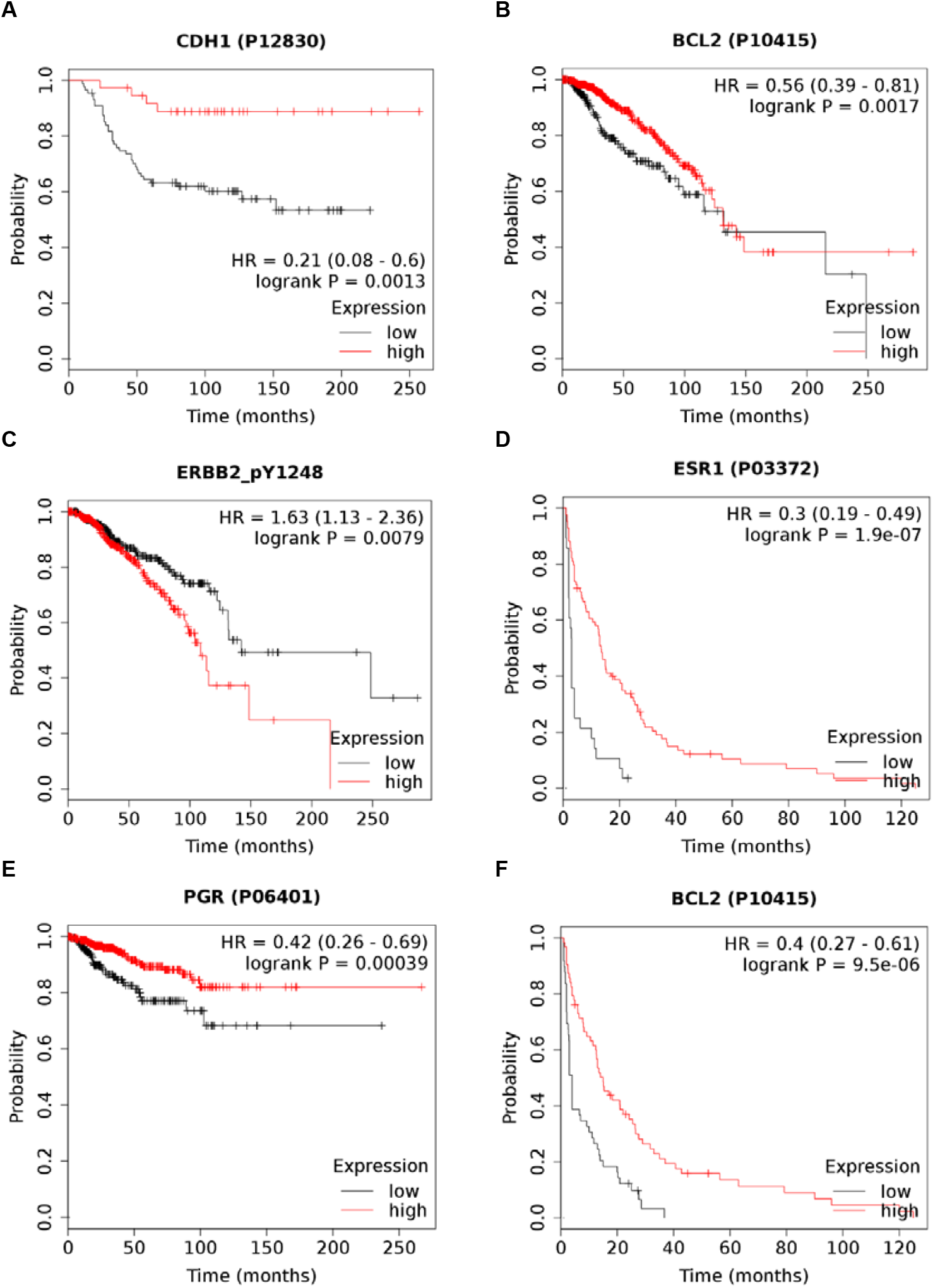
Survival outcome differences in patients with different expression levels of protein biomarkers. Kaplan-Meier plots of overall survival by CDH1 (E-cadherin) **(A)**, apoptosis regulator BCL2 **(B)**, and tyrosine 1248-phosphorylated HER2 **(C)**. Kaplan-Meier plots of relapse-free survival for estrogen receptor 1 **(D**), progesterone receptor **(E)** and BCL2 **(F)** in breast cancer patients. Note the different survival characteristics of the different datasets.

## DISCUSSION

A major advance of proteomic technologies lies in their ability to simultaneously measure multiple biomarkers from a single clinical specimen. Here, we collected four independent breast cancer proteomic cohorts and validated established and new biomarker candidates.

Despite the quantitative and multiplexing limitations of immunohistochemical analysis, in clinical practice, it is still the gold standard. We compared the efficiency of various proteomic techniques to determine routinely measured breast cancer biomarkers, including ESR1, PGR, HER2, and MKI67. In this analysis, both the RPPA and LC-MS/MS method results were highly correlated with IHC results and thus can be utilized to determine receptor status in breast cancer patients. Unfortunately, we did not have all markers for the same patients, and the results achieved for individual genes can only suggest that proteomic technologies will also be capable of performing molecular stratification in the future, enabling the discrimination of breast cancer subtypes.

Estrogen receptor is a pioneer cancer biomarker, and classifying breast tumors based on hormone receptor status has been utilized in routine clinical practice for over four decades (24). ESR1 positivity and PGR positivity are associated with better survival outcomes than negative ESR1/PGR status. In addition to clinicopathological prognostication, the main medical application of these receptors is selecting patients for endocrine therapy (25).

MKI67 is a protein not expressed in G0 phase, and thus, it is a perfect marker for determining the proportion of dividing cells. MKI67 expression is correlated with outcome, and high MKI67 expression is associated with poor prognosis, which has been validated in a meta-analysis involving over 64 thousand breast cancer patients (26). Immunohistochemical staining of MKI67 alone can also pinpoint low-risk breast cancers with the same reliability as genomic markers (27).

Evaluation of HER2 (ERBB2, neu) status has also been routinely used in breast cancer molecular diagnostics since the end of the 1990s. Analysis of large cohorts of patients found that HER2 overexpression is associated with unfavorable prognosis and poor response to chemotherapy (28). The clinical introduction of anti-HER2 therapies (i.e., trastuzumab, pertuzumab) in combination with chemotherapy in patients who have HER2-positive cancer results in exceptional survival advantages. As a result, HER2-positive patients have a better outlook than HER2-negative patients (29). Today, tumors with even 1% positivity are eligible for anti-HER2 therapy (30).

Triple-negative breast cancer (TNBC) is diagnosed in cases where tumors are negative for ESR1, PGR, and HER2. In these breast tumors, the immunohistochemical measurement of basal markers (cytokeratin 5/6, EGFR), claudins (CLD3/4/7), cadherins (CDH1, CDH3), stem cell markers (CD44/CD24, ALDH1), apoptosis markers (BCL2, TP53), a transcription marker (YB-1) and urokinase-type plasminogen activator (uPA)/plasminogen activator inhibitor-1 (PAI-1) have also been suggested for advanced stratification (16, 17, 31, 32).

We assessed the prognostic power of a selected set of proteins, including ESR1, PGR, HER2, cytokeratins, claudins, E-cadherin (33) and EGFR, in the datasets included in the present study. Overall, we uncovered that 33 proteins had a significant correlation with prognosis. In the case of FDA-approved protein biomarkers, the expression of estrogen and progesterone receptors is correlated with favorable relapse-free survival. High expression levels of phosphorylated HER2 protein measured by RPPA were linked with worse overall survival than low expression levels; these findings are in line with the previous study by Hayashi et al. on the same protein (34).

High expression of the antiapoptotic Bcl-2 and the adhesion marker E-cadherin was related to longer relapse-free survival than low expression in at least two independent datasets. Bcl-2 overexpression was revealed in other cancers and was linked to cancer initiation and progression, and higher expression positively correlated with favorable patient outcomes in hormone receptor-positive breast tumors (35, 36). Loss of E-cadherin expression is frequently represented in invasive lobular breast carcinoma, which is three times more likely to metastasize (37).

Interestingly, some of the genes, including PGR and E-cadherin, display inverse correlations with survival when assessing the link to survival in different patient cohorts. Here, we have to mention some limitations of our analysis that might lie behind these discrepancies. A major constraint is that only 20% of the proteins were determined in at least three platforms. This means that the evaluation of further databases will be needed to perform a comprehensive validation of all potential biomarker candidates. Another shortcoming of the investigated datasets is the rather low proportion of events (in the case of the TCGA dataset) and the short follow-up time (DeMarchi dataset). A future large-scale proteomic database with long follow-up and uniform protein level determination using a single method could provide more reliable data for a similar analysis.

In summary, we successfully integrated four distinct breast cancer proteomic datasets containing tumor and normal samples. A significant correlation was observed between marker levels detected by proteomic technologies and those detected by immunohistochemistry results. We validated prognostic and predictive breast cancer biomarkers and compared the efficiency of different proteome analysis techniques. The entire database is integrated into our online tool, providing an opportunity to validate our findings and to identify and rank new survival-associated biomarker candidates using multiple independent cohorts of breast cancer.

## Data Availability

Data sharing is not applicable to this article as no new data were created or analyzed in this study, however, we expanded our established web tool to validate survival-associated biomarkers to include the proteomic datasets analyzed in this study (https://kmplot.com/analysis/).

## AUTHOR CONTRIBUTIONS

Concept and design: BG; Database setup: OA, BG; Analysis of data: OA, BG, AL; Data interpretation: OA, BG; Draft manuscript: OA, BG. All authors provided final approval of the manuscript.

## ACKNOWLEDGEMENTS AND FUNDINGS

The study was supported by the 2018-2.1.17-TET-KR-00001, 2018-1.3.1-VKE-2018-00032 and KH-129581 grants of the National Research, Development and Innovation Office, Hungary. The use of the computational infrastructure of Pázmány Péter University, provided within the National Bionics Program, is gratefully acknowledged. The authors acknowledge the support of ELIXIR Hungary (www.elixir-hungary.org).

## SUPPLEMENTARY MATERIAL

Supplemental table 1: Protein list

Supplemental table 2: Protein biomarkers

